# AI Algorithm Deployment and Utilization: Strategies to Facilitate Clinical Workflow Implementation and Integration

**DOI:** 10.1101/2023.09.19.23295729

**Authors:** Barbaros Selnur Erdal, Alexis Laugerette, Kimberly H. Fair, Mathis Zimmermann, Mutlu Demirer, Vikash Gupta, Thomas P. O’Donnell, Richard D. White

## Abstract

Based on past experiences of the Center for Augmented Intelligence in Imaging (CAII) [Department of Radiology, Mayo Clinic Florida], depending on the project, 10 to 20 months has typically been required to realize the successful creation (data curation and algorithm development), and utilization (integration, testing, and operationalization) of an AI algorithm [Figure 1].

Figure 1:
AI algorithm evolution typically requires 10 to 20 months, consisting of four consecutive phases: 1. data identification and extraction; 2. data cleansing and labeling; 3. algorithm development with training and tuning; and 4. Implementation and integration with testing and operationalization.

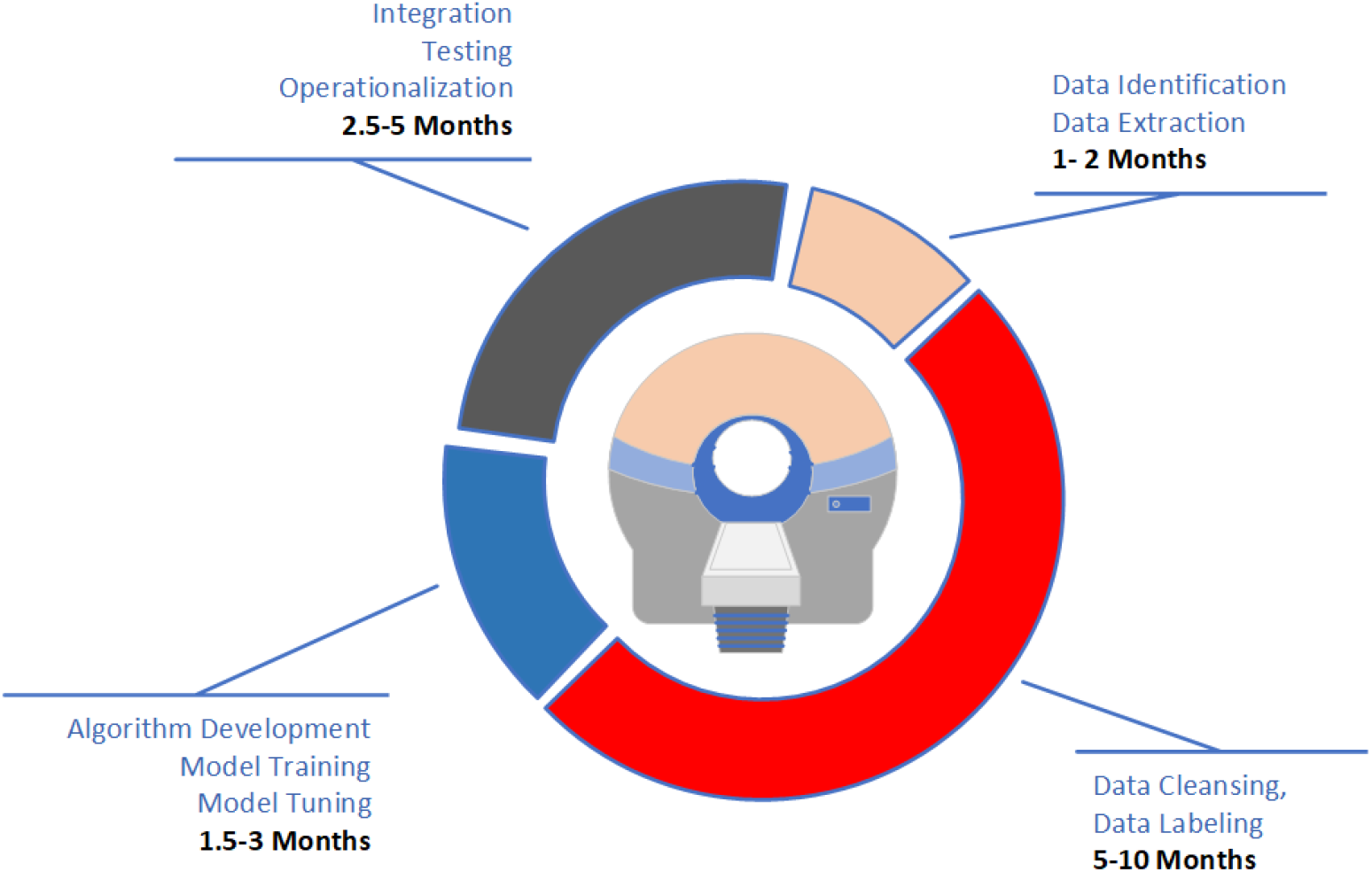

This manuscript delineates the related challenges and opportunities for greater efficiency in completing the clinical workflow implementation and integration of an AI algorithm. Strategies exploiting conventional data standards in facilitating the completion of such deployment and utilization goals within the operations of a busy Radiology practice are described. Methodologies and techniques employed during this initial phase of the CAII-Siemens D&A AI collaboration to address the previously mentioned challenges and opportunities are depicted with use-case examples.

## Background

Data contributing to the evolution of an AI algorithm may potentially come from multiple sources and change over time. Accordingly, the supporting framework must be able to: 1. ingest the output from a previous module; 2. provide physically meaningful data augmentation; 3. expand to incorporate new image and non-image data types; and 4. recycle previously curated data and developed algorithms.

Regardless of whether they are vendor-based or locally developed, data-flow leading to the creation of an AI algorithm or to AI-algorithm implementation and integration may have to contend with complex demands. These demands are best met via an IT architecture facilitating interoperability by incorporation of common data standards (e.g., HL7 [1] and DICOM [2]) along with IHE [3] profiles that describe how these standards can be used to interconnect applications for efficient regular clinical workflow [Figure 2].

**Figure 2:**
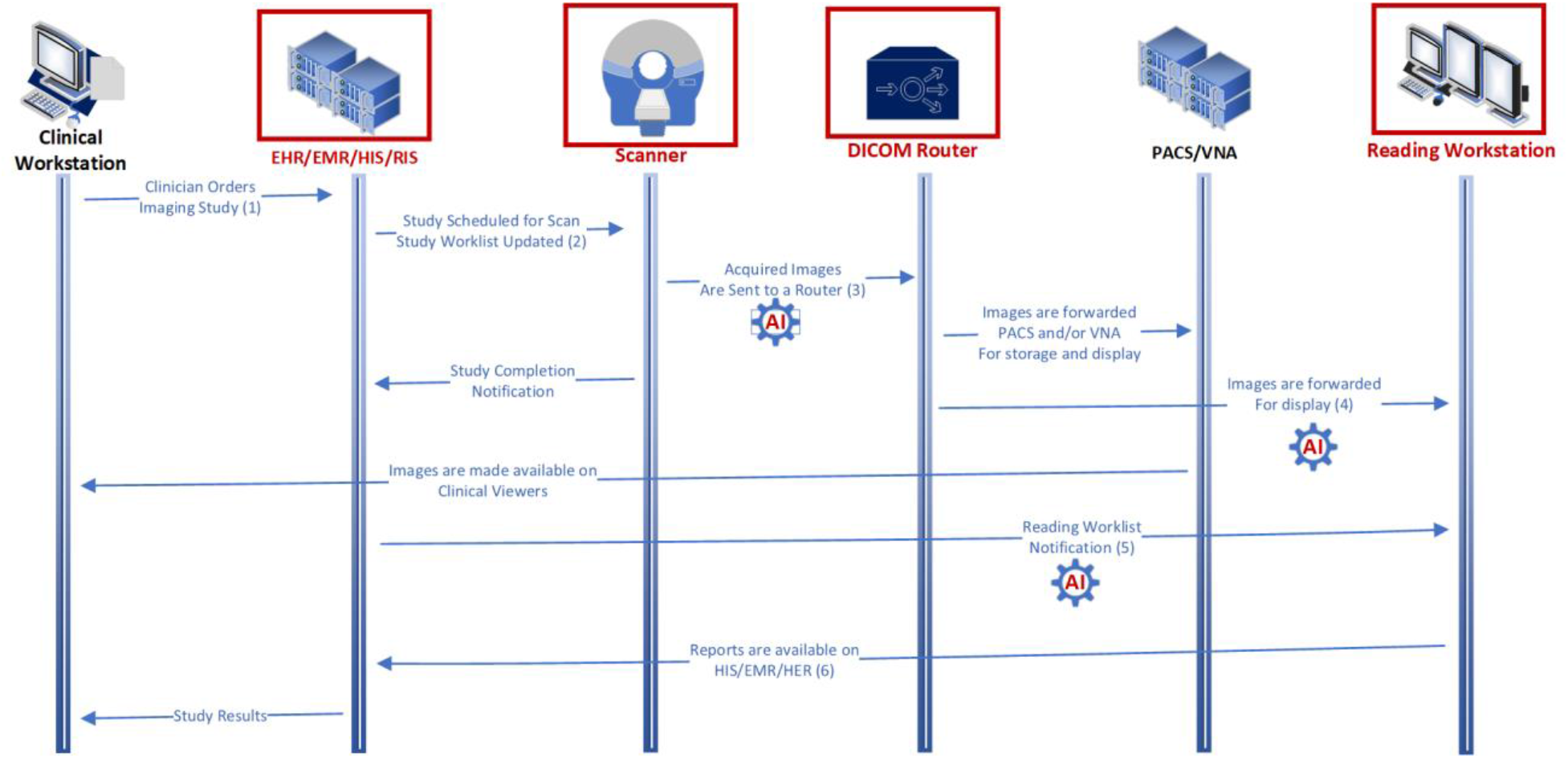
Basic Radiology workflow, modeled after IHE Scheduled Workflow. Shown are: 1. an order being generated; 2. image-data being acquired during patient scanning; 3. produced images being evaluated by a radiologist; and 4. a report being generated by the image interpreter and sent back to the clinician for review. AI-sprocket symbols indicate examples of AI use-cases.

Understanding the interoperability potential within a given workflow pattern is the first step in considering AI integration touchpoints. The types of workflows are described in the IHE AI in Imaging White Paper [4], which references AI Workflow for Imaging (AIW-I) [5] and AI Results (AIR) [6], two interoperability profiles (including boundaries and transactions) tailored for AI applications.

Examples of AI application to clinical workflows are varied [e.g., Figure 2 illustrated by the AI-sprocket symbol] and relate uniquely to AIW-I and AIR [Table 1].

**Table 1:** IHE Profiles and Workflow Steps Covered in Initial Phase of CAII-Siemens D&A AI collaboration.

| Steps | Common Workflow Theme | AIW-I and AIR Described Boundaries | Covered Steps |
| --- | --- | --- | --- |
| 1 | When a clinician orders an imaging examination in the HIS/RIS, they may be guided by a Clinical Decision Support System (CDSS) to ensure its appropriateness. Depending on the clinical setting, the order may contain a clinical-status priority code (e.g., "stat") | AI applications (CDSS) may make recommendations as to the types of procedures that should be ordered, based on the patient's condition and record. |  |
| 2 | Once the patient examination is scheduled for a date and location, an entry is created on the "study worklist" of the scanner (or another imaging device). In some instances, an entry is also created on a "protocoling worklist," where a radiologist determines the specific imaging techniques to be used (e.g., scanning details, contrast-agent type/amount/administration route) during the diagnostic imaging study or image-directed procedure. | AI applications may make recommendations on the type of protocol to be used on the scanner. |  |
| 3 | Once the examination is completed, images are reconstructed into a human-interpretable format and sent to a DICOM-router to be forwarded to the appropriate destinations, including a PACS and/or VNA for management or storage. Once the organized images (original and/or post-processed) are ready to be evaluated by the radiologist, the examination description appears on the radiologist's "reading worklist". | AI applications may (automatically) post-process images, identify QA issues prior to the patient leaving the department, and prepare classifications and segmentations in advance of the radiologist's evaluation. AI results may also influence the reading priority of the exam. | X |
| 4 | Radiologists assess the examination images on their diagnostic viewer and dictate their interpretation (typically into a voice recognition-based reporting system). | AI applications may provide insights to be included alongside the images in the radiologist's display. | X |
| 5 | The dictated report is sent to the HIS/RIS. If actionable critical and/or non-critical findings are identified, radiologists may invoke additional workflows to alert the ordering clinician and issue the final examination report. | The workflow may include emergent insights provided by AI applications, for consideration of the ordering physician. | X |
| 6 | Final examination reports become available in the HIS/EHR, along with the images in the PACS or clinical viewers. | Pre-populate the radiologist's report with draft insights to be considered by the radiologist. |  |

## Methods

### General Principles

Several AI applications were implemented for research purposes in the secure CAII IT test environment [7].

The Siemens Healthineers AI Rad Companion (AIRC) Chest X-Ray application was deployed in the cloud and integrated into CAII research systems as per the workflow [Figure 3].

**Figure 3:**
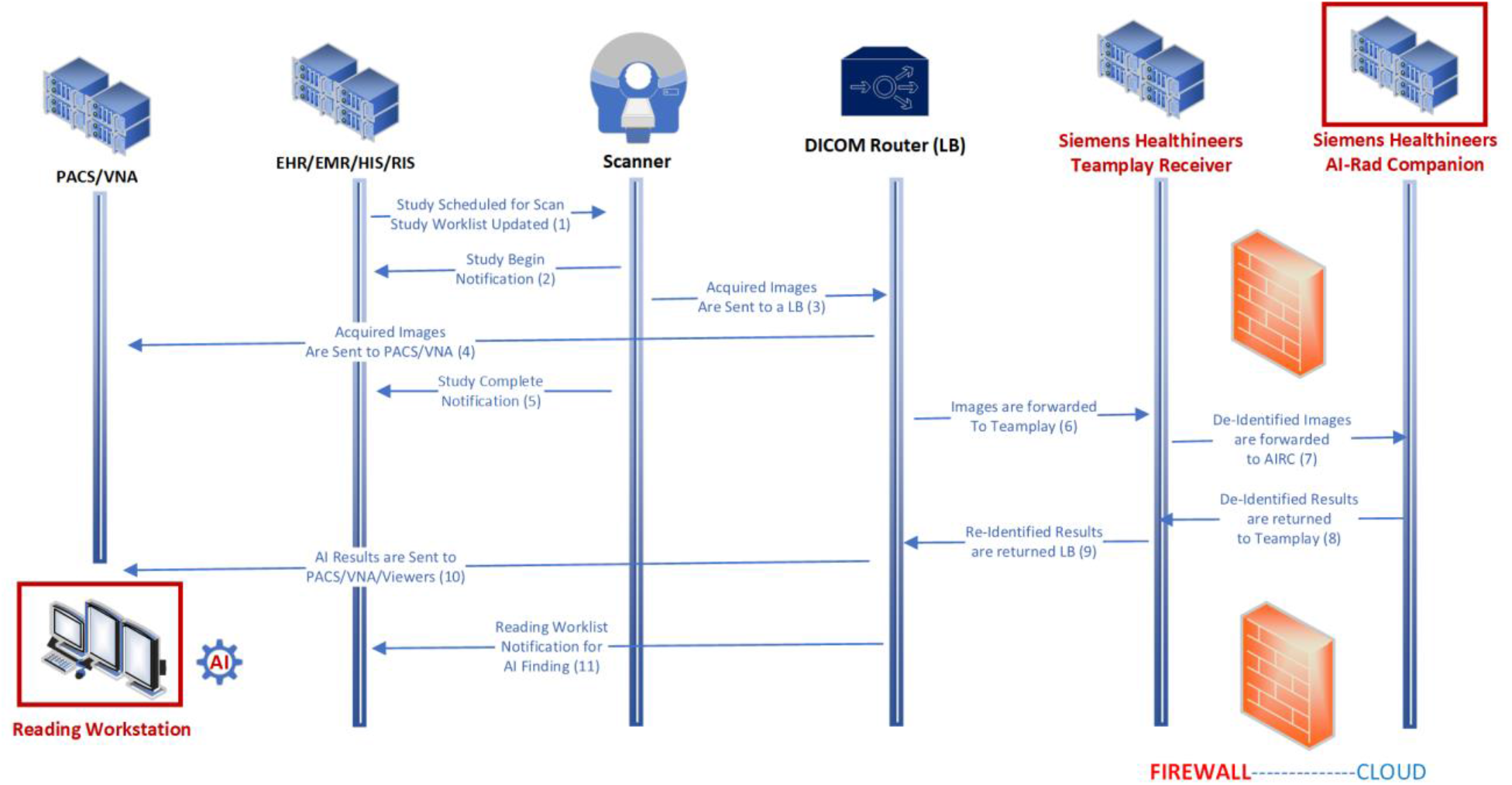
Workflow Steps Other AIRC applications (e.g., Chest CT) were made available using the same infrastructure.

Locally developed AI applications (e.g., MRI-safety screening algorithm for leadless electronic implanted device detection and identification on Chest X-ray) were similarly deployed while running on-premises [7], thereby obviating the need for the DICOM gateway Siemens Healthineers Teamplay Receiver (TPR). With several locally developed algorithms already deployed and operational, coordinated integration with a vendor-based solution maintained existing workflows and interoperability.

### Implemented Workflow for Outside Vendor Integrations

The workflow steps followed by a given study within the CAII infrastructure includes:

1. An exam is scheduled when Clinical Team orders an imaging study. This triggers an update on Study Worklist (e.g., Modality Worklist)
2. When Imaging Team works on scheduled exam, a “begin exam” message is sent to HIS/RIS (Epic).
3. Acquired images are sent to DICOM Router used locally (i.e., Laurel Bridge (LB)).
4. LB sends acquired images to combined clinical and non-clinical PACS/VNA.
5. When image acquisition is fully completed, a “study complete” message is sent to HIS/RIS.
6. Applicable AI workflows are triggered: Based on exam codes, LB sends a copy of acquired images to Siemens Healthineers TPR, a DICOM gateway application. Considered exam codes were for the following procedures: DX Chest 1 View, DX Chest 2 Views, DX Chest AP or PA.
7. TPR (operating on-premises and managed by local Imaging Informatics Team Members) de-identifies the images and forwards them to Siemens Healthineers AIRC (AIRC).
8. AIRC (operating on cloud) runs inference on forwarded images and returns DICOM results to TPR. These results follow formats recommended within IHE AIR profile (e.g., DICOM SR TID1500).
9. TPR re-identifies produced DICOM results and returns them to LB.
10. LB forwards results to a research/non-clinical PACS/VNA (replica of institution’s clinical PACS i.e., Visage) as well as to the CAII adjudication viewer DICOM node. The CAII viewer offers capabilities to display AI results along with original images, and to interact with these results (e.g., to edit them).
11. A custom program executed on LB (written in C#) creates HL7 message (based on DICOM SR content) to trigger an event on a test instance (non-clinical) of institution’s HIS/RIS (i.e., Epic Radiant), if a given result is expected to produce a priority message (e.g., Radiology reading worklists can be re-sorted or color-coded using this mechanism).

This approach can be summarized in the following pseudo-code:

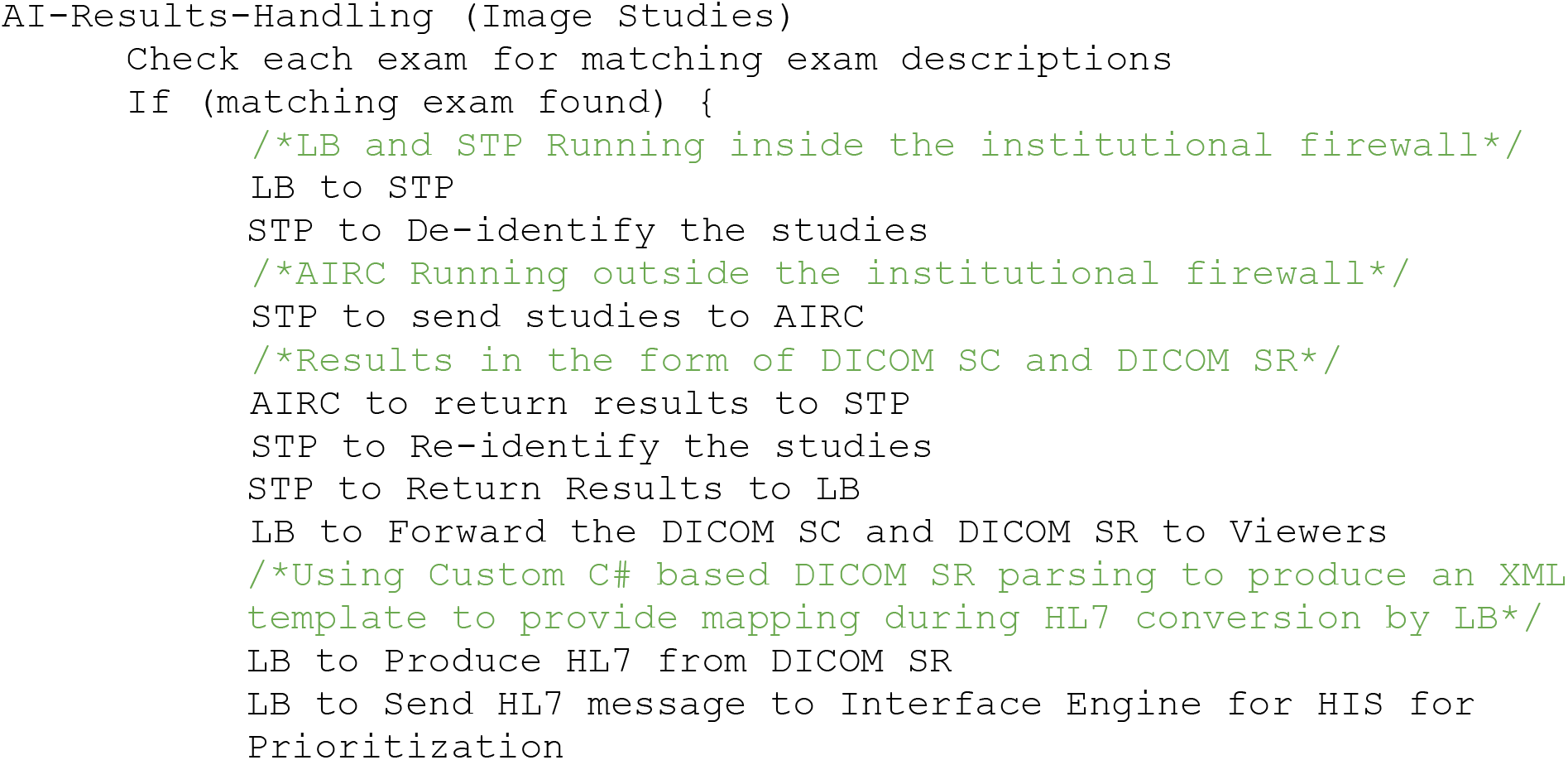

### Worklist Management

The worklist re-prioritization was achieved by parsing the resulting DICOM SR content and generating a simple HL7 message using the following example template (i.e., pneumothorax detection), filling the necessary fields either by rule-based hardcoding or by fetching content from DICOM metadata:

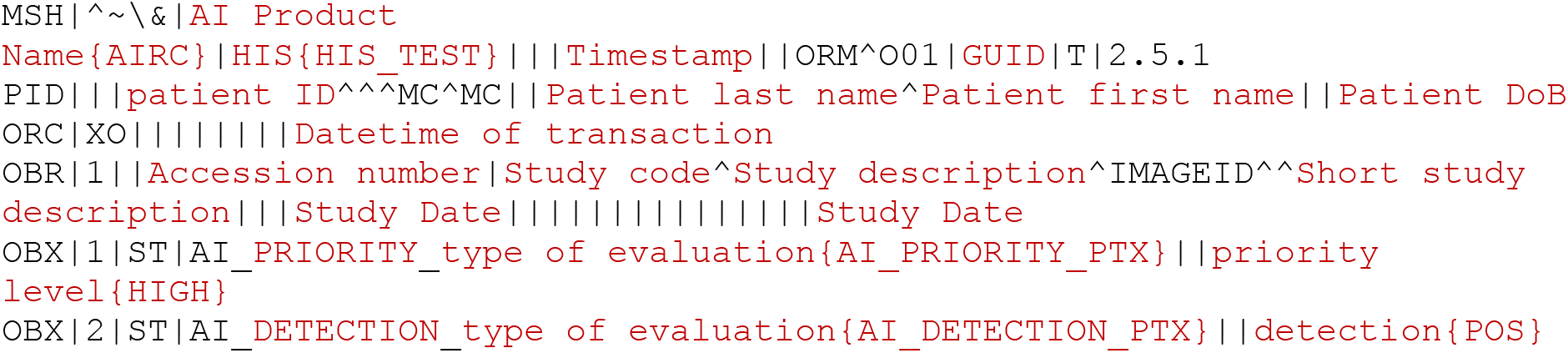

### Usage Ramp-Up and First-Level Integration Test

Although the focus of the project was on Chest X-ray use, several applications from the AIRC collection were also used to establish the feasibility of integration of several applications into a single viewing environment based on a variety of standard display and structured formats.

Such was the case with AIRC Chest CT, which was successfully and easily integrated into the existing infrastructure based on the locally developed CAII Viewer [7] using existing DICOM Secondary Capture and DICOM SR TID1500 outputs, the last providing more interaction capabilities with the AI results [Figure 4]. Connecting different AIRC applications to the same infrastructure was a helpful step in assessing the robustness of the established connections.

**Figure 4:**
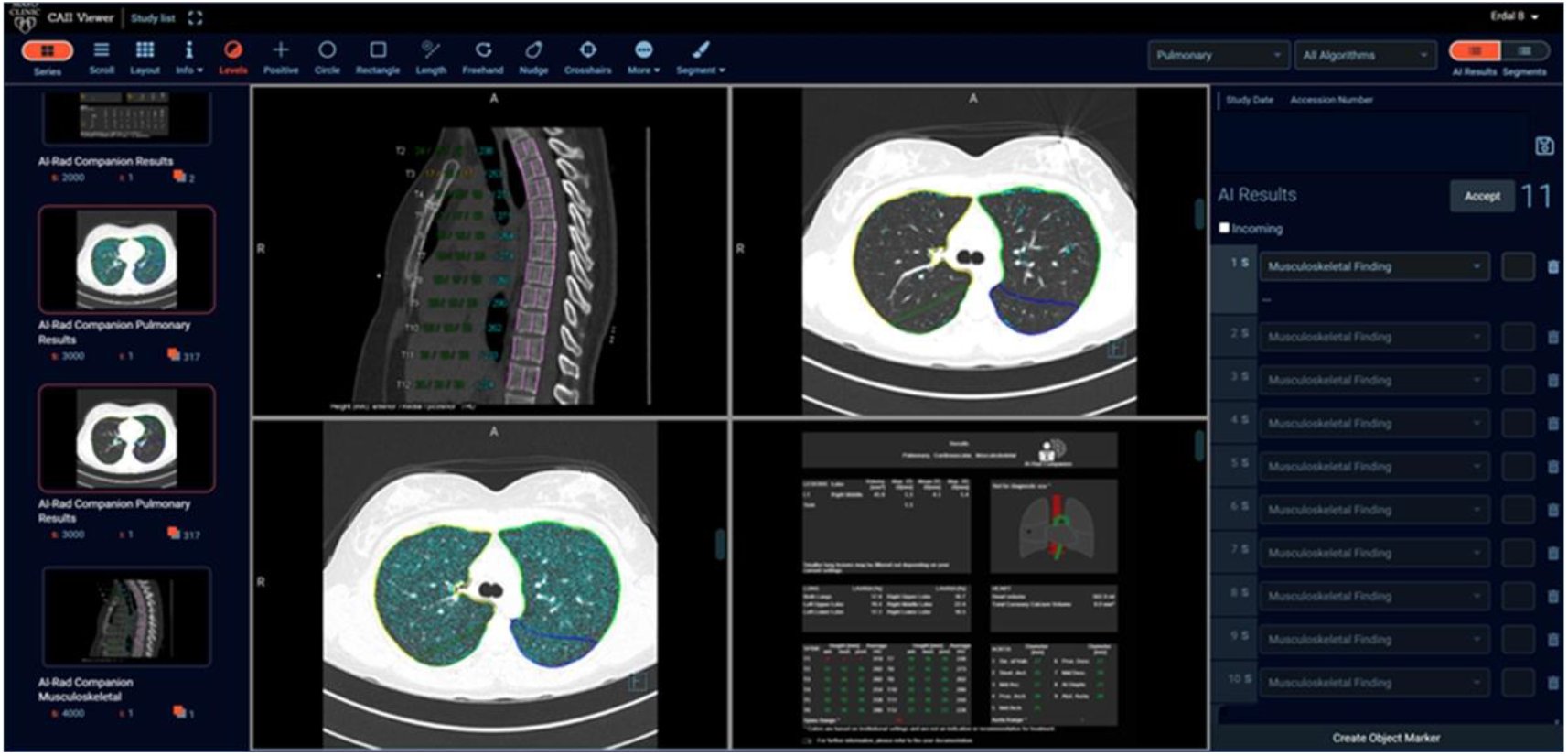
In addition to cross-platform deployment and display capabilities, common standards-based algorithm results enable user feedback capabilities across platforms supporting these formats, as illustrated on the locally developed CAII Viewer and AIRC (DICOM Secondary Capture and DICOM SR, on the right-hand-side, of results).

## Results

### AI Processing at Scale for Enterprise

The Siemens Healthineers AIRC Chest X-Ray algorithm was deployed within the CAII test environment using the previously described framework and methods. AIRC Chest X-ray was activated for over five months (January 1, 2023 to May 31, 2023), during which imaging studies were sent to AIRC from 20 different digital X-ray machines operating with the Mayo Clinic Florida Department of Radiology and representing a range of manufacturers (i.e., Siemens Healthineers 79%, Samsung 16%, Carestream 5%).

During the trial period, a total of 22,885 Chest X-ray images (approximately 17,000 individual exams) were received by AIRC. The discrepancy reflects the fact that an exam may contain more than one image matching input image requirements of AIRC (e.g., a Chest X-Ray study with several retake images, with each image suitable for AIRC processing); therefore, the number of individual images analyzed by AIRC exceeds the count of individual exams over the same period.

In the trial, AIRC processing of an image could potentially end in one of three results:

- Successful Processing: The algorithm ran successfully, and an inference was produced.
- Failed: There was an error in algorithm execution, and no inference was produced.
- Unsupported Input: The received image was not suitable for processing, and consequently, no result was produced.

The details of processing activity (Figure 5) and results statistics (Table 2) follow:

**Table 2:** Processing Statistics Achieved with AIRC Chest X-Ray in CAII Infrastructure Over Trial Period.

| Result of Processing | Count | % of Total Images Received by AIRC | Average # Images Received per Day | Comments |
| --- | --- | --- | --- | --- |
| Processed Successfully | 22,382 | 97.80% | 154/day | Results available within 3 minutes after image reception by TPR |
| Failed Processing (No Results Created) | 2 | 0.01% | <<1/day |  |
| Unsupported Input (No Results Created) | 501 | 2.19% | 3/day | Reasons include unsupported image types, patient demographics (age), other check on DICOM attributes |
| Total Number of Images Received by AIRC | 22,885 | 100% | 158/day | Corresponding to 113 individual exams/day |

**Figure 5:**
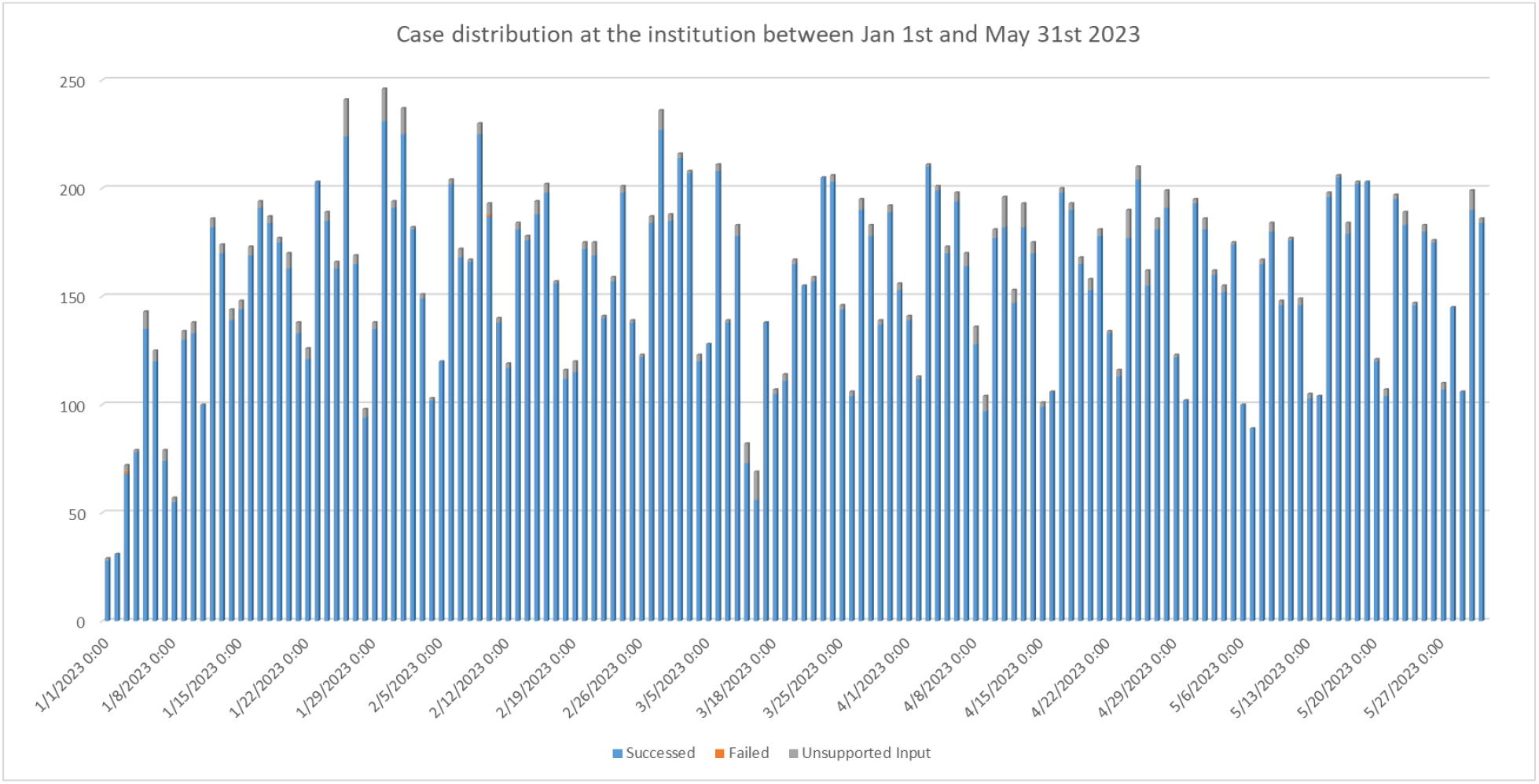
Distribution of Daily Processing Activity Over Trial Period.

In this project, we showed that by using a common standards-based architecture and workflow the ability to deploy algorithms, whether vendor-based (commercial products or prototypes) or locally developed [7], can carry significant advantages. This approach enables support of complex operational requirements and allows both types of solutions to coexist in the same environment. As a result, end-users can benefit from concurrent AI support of multitude of tasks, including the adjudication of AI results (e.g., for algorithm continuous learning, data collection, product improvement) to more operational uses of AI results (e.g., for reading worklist management).

### Worklist Management

We demonstrated that worklist re-prioritization was possible using standard-based outputs (DICOM SR) and appropriate middleware systems (e.g., a DICOM router and custom parsing logic for HL7 message generation). Worklist re-prioritization was successfully observed for patients with AI-suspected pneumothorax and could be visualized (Figure 6) using the CAII infrastructure.

**Figure 6:**
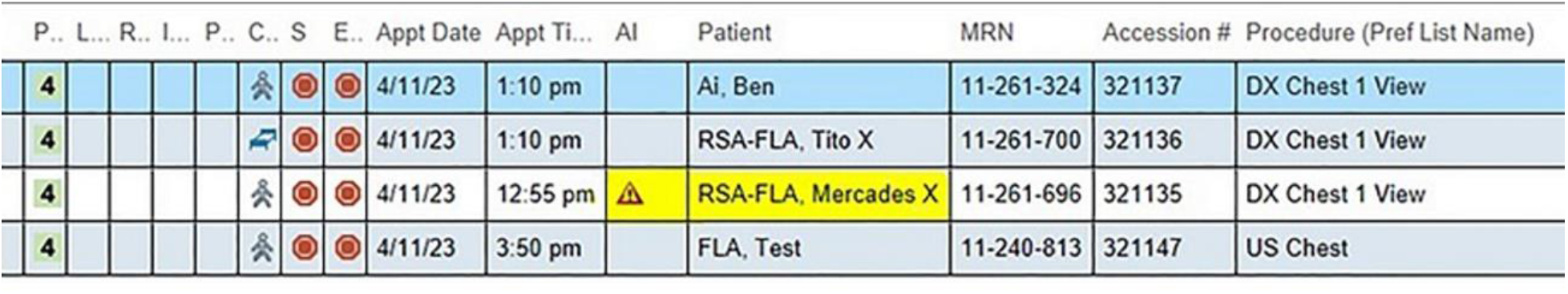
An HIS Interface Example Demonstrating Reading Worklist Prioritization Based on AI Findings.

### Statistical View of AIRC-Processed Images

Over the trial period (January 1, 2023 to May 31, 2023), from 22,382 successfully processed images, AIRC produced 33,090 automated image annotations (detected findings or no findings detected). Close to 80% of the annotations produced by AIRC related to a detected disease; the remaining 20% were for images flagged with “none of the findings detected” by AIRC.

A detailed breakdown of this data (Table 3), and a corresponding diagram using a Sankey diagram (Figure 7) follow. Consolidation was the most frequently detected abnormality, followed by no detected abnormality.

**Table 3:** Diseases Detected by AIRC Over Trial Period. The first column shows annotations produced by AIRC (e.g., one analyzed image can contain more than one disease annotation at one location, as well as different diseases at different locations). The last two columns indicate the frequency of unique images identified as having at least one annotation per disease (an image can have one or more diseases, allowing an image to appear in more than one category).

| **Disease Annotations** | **# of Annotations** | **% of Annotations** | **# of Unique Images with Disease** | **% of Unique Images with Disease** |
| --- | --- | --- | --- | --- |
| Consolidation | 21,844 | 66.0% | 13,469 | 55.8% |
| Atelectasis | 1,360 | 4.1% | 1,199 | 5.0% |
| Lesion | 1,287 | 3.9% | 1,098 | 4.5% |
| Pleural Effusion | 1,076 | 3.3% | 889 | 3.7% |
| Pneumothorax | 874 | 2.6% | 836 | 3.5% |
| No detected finding<br>(None of 5 above) | 6,649 | 20.1% | 6,649 | 27.5% |
| Total | 33,090 | 100.0% | 24,140 | 100.0% |

**Figure 7:**
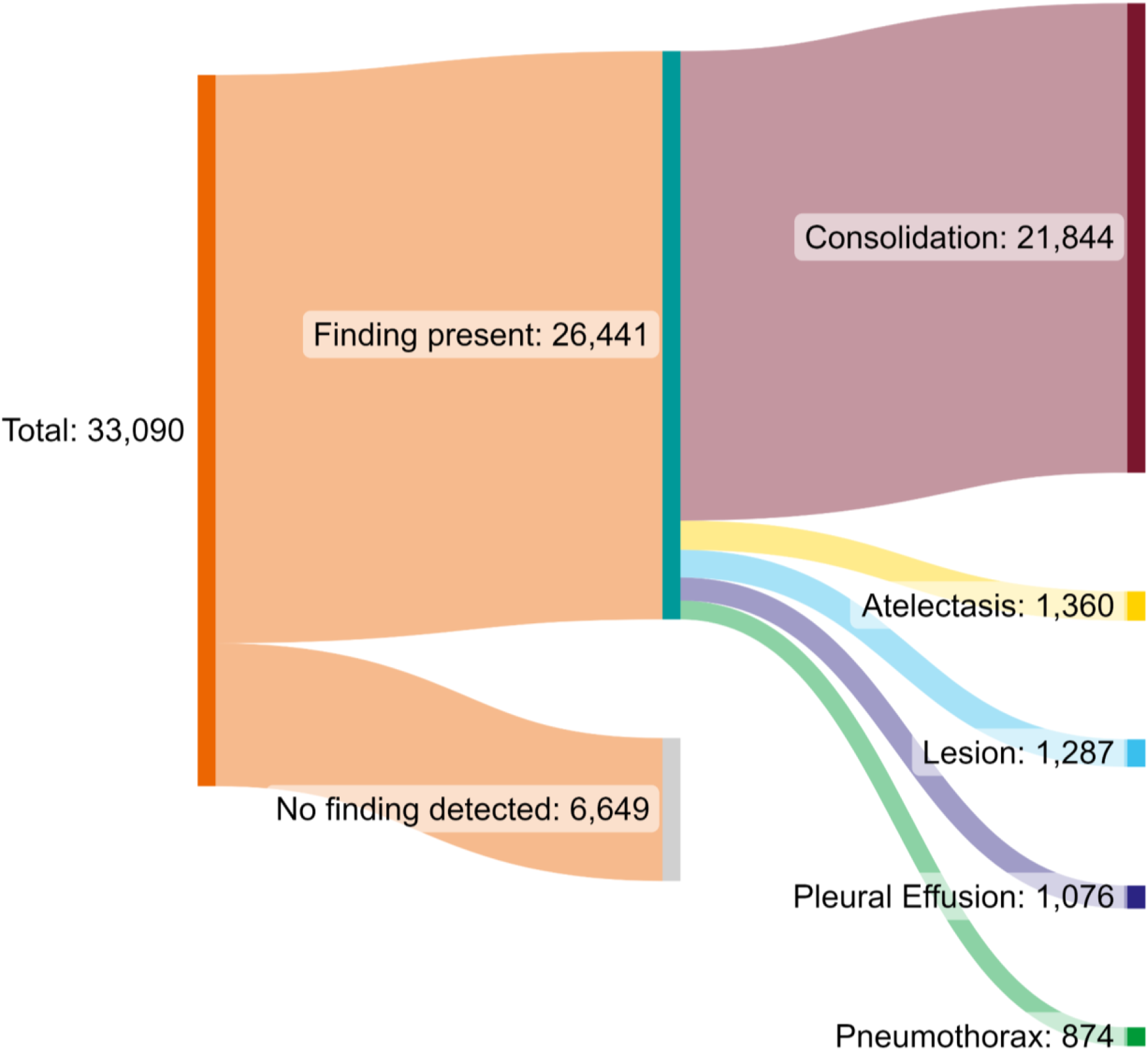
Sankey Diagram Representing the Breakdown of Disease Annotations Generated by AIRC Chest X-Ray During Trial Period.

Comparison against radiologist “gold standard” reporting, to draw conclusions on the AI algorithm accuracy (i.e., correct diagnosis) and utility (e.g., for use-cases like worklist management and pneumothorax flagging), was not evaluated, as this project was focused on workflow integration and informatics/deployments aspects rather than on algorithm performance. Accordingly, exam-based clinical conclusions cannot be made as only statistics per image (one exam potentially having more than one image) have been computed. Similarly, images identified as “no finding detected” may not be considered to be “normals”. “No finding detected” only indicates the perceived absence of the 5 diseases under consideration (i.e., pneumothorax, lesion, atelectasis, consolidation, pleural effusion); other findings not addressed by the algorithm(s) running may actually be present in the images. However, such investigations may be performed in the future to expand on this project.

### Viewing Infrastructure: Interacting with AI Inference Results Based on Standardized Outputs

The CAII Viewer employed in this project successfully demonstrated that standards-based AI-result reporting offers the ability to capture feedback from user adjudication (accept, reject, edit) (Figure 8 and Figure 9a-c) from a single source, but more importantly from multiple from different sources (e.g., different algorithms, whether vendor-based or locally developed) in a single viewing environment. For example, this was shown using the AIRC Chest X-Ray results (vendor product) and a locally developed leadless implanted electronic device detection and localization algorithm for determination of MRI safety based on Chest X-Rays; both algorithms were able to run in parallel on the same datasets, and inference results from both (transmitted in the form of DICOM SR TID1500) could be visualized simultaneously in the CAII Viewer (Figure 10).

**Figure 8:**
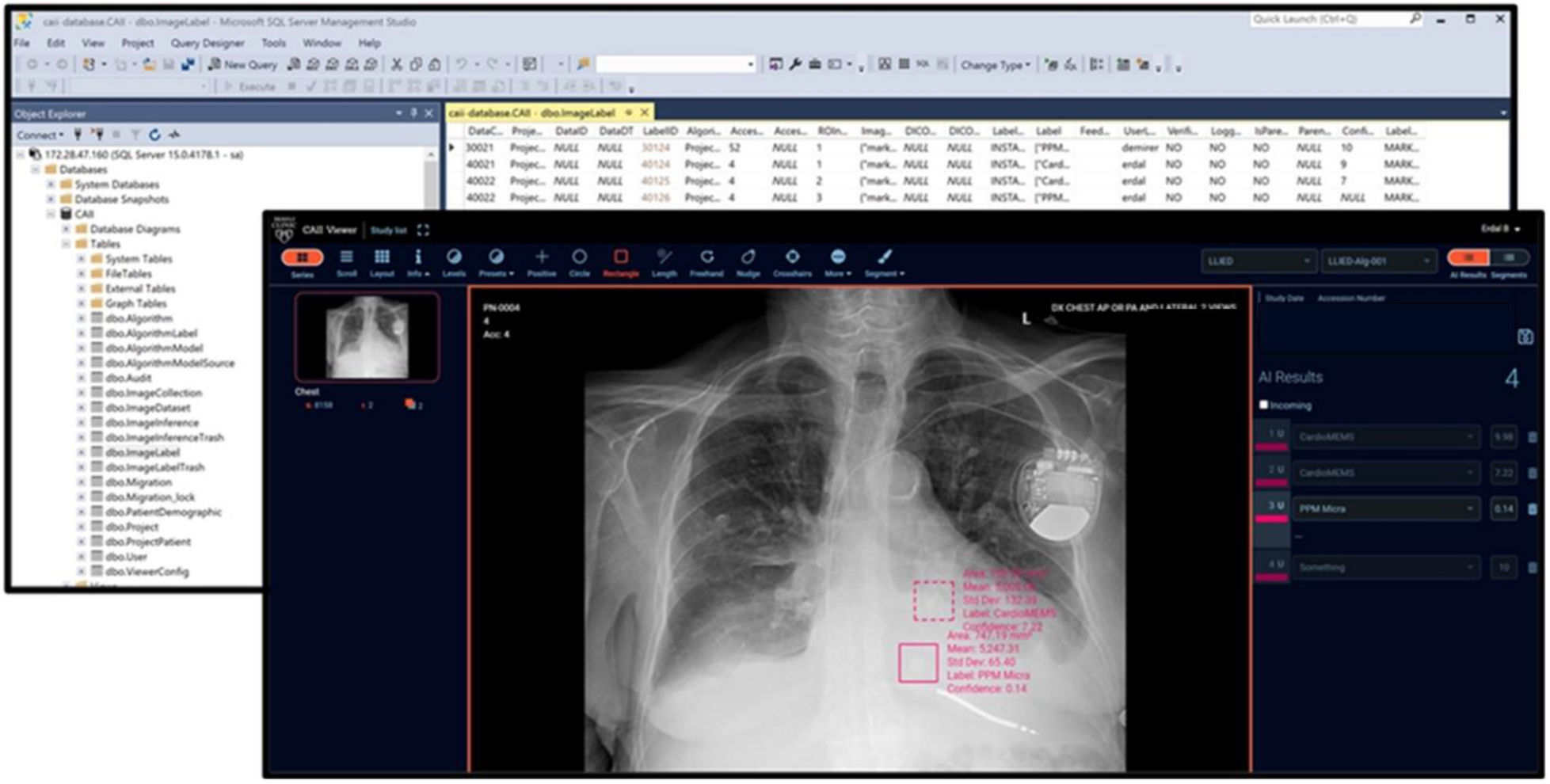
Being able to capture standards-based user feedback enables database-driven algorithm development and enhancement, as well as easy and structured data capture. This is illustrated with a locally developed algorithm for leadless implanted electronic device algorithm for determination of MRI safety based on Chest X-Rays [7]. This algorithm is trained to recognize potentially MRI-unsafe devices using two cascading AI models (the results of the first model for detection based on a fast R-CNN then feed into the second model for identification based on a multi-class CNN).

**Figure 9a:**
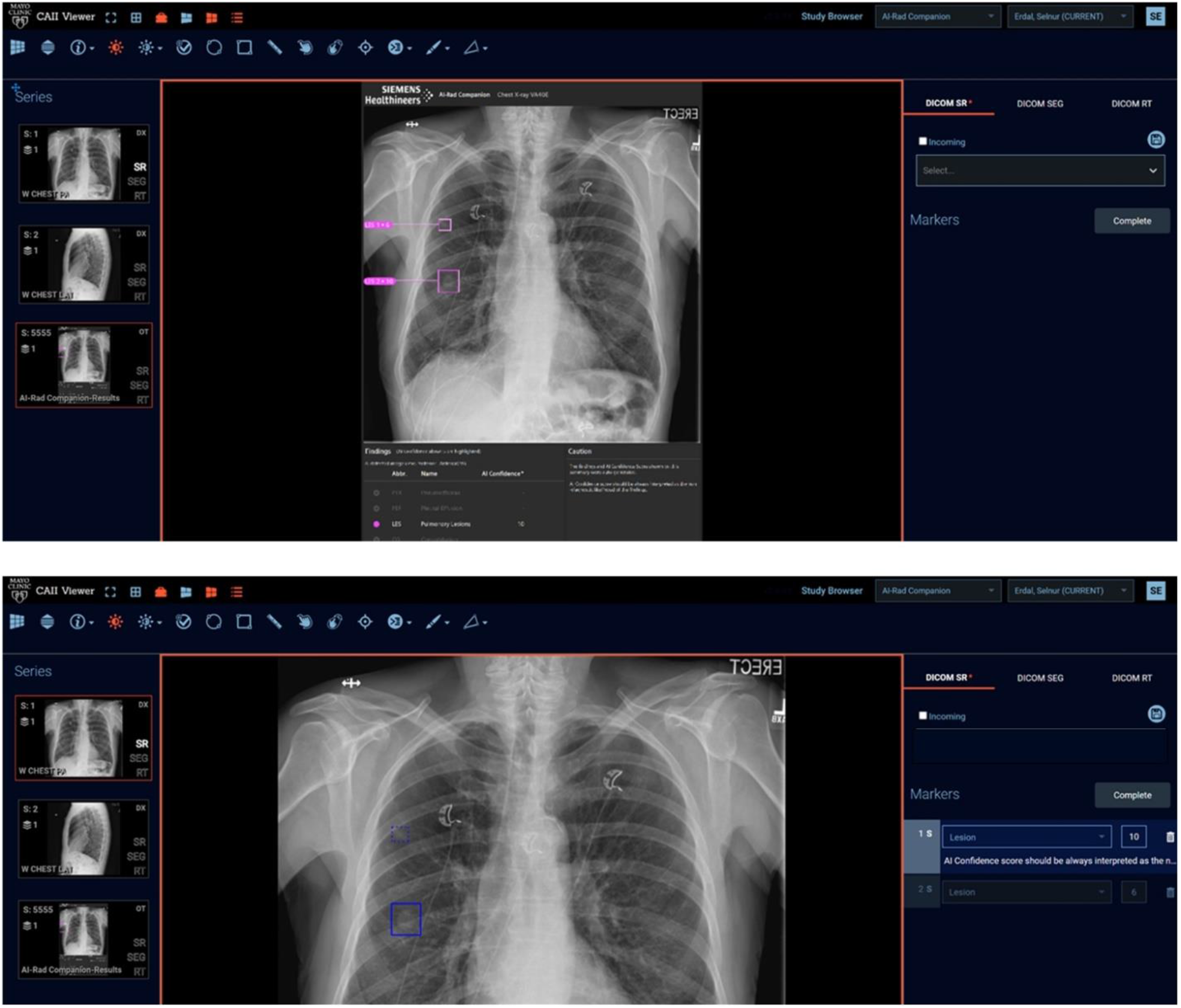
AIRC Chest X-ray results showing a subtle lung lesion as displayed in the locally developed CAII Viewer [7]. The CAII Viewer can display a wide range of DICOM formats, including non-editable Secondary Captures (as shown here), as well as DICOM SR outputs (left-hand-side menu) for more control by the end-user.

**Figure 9b:**
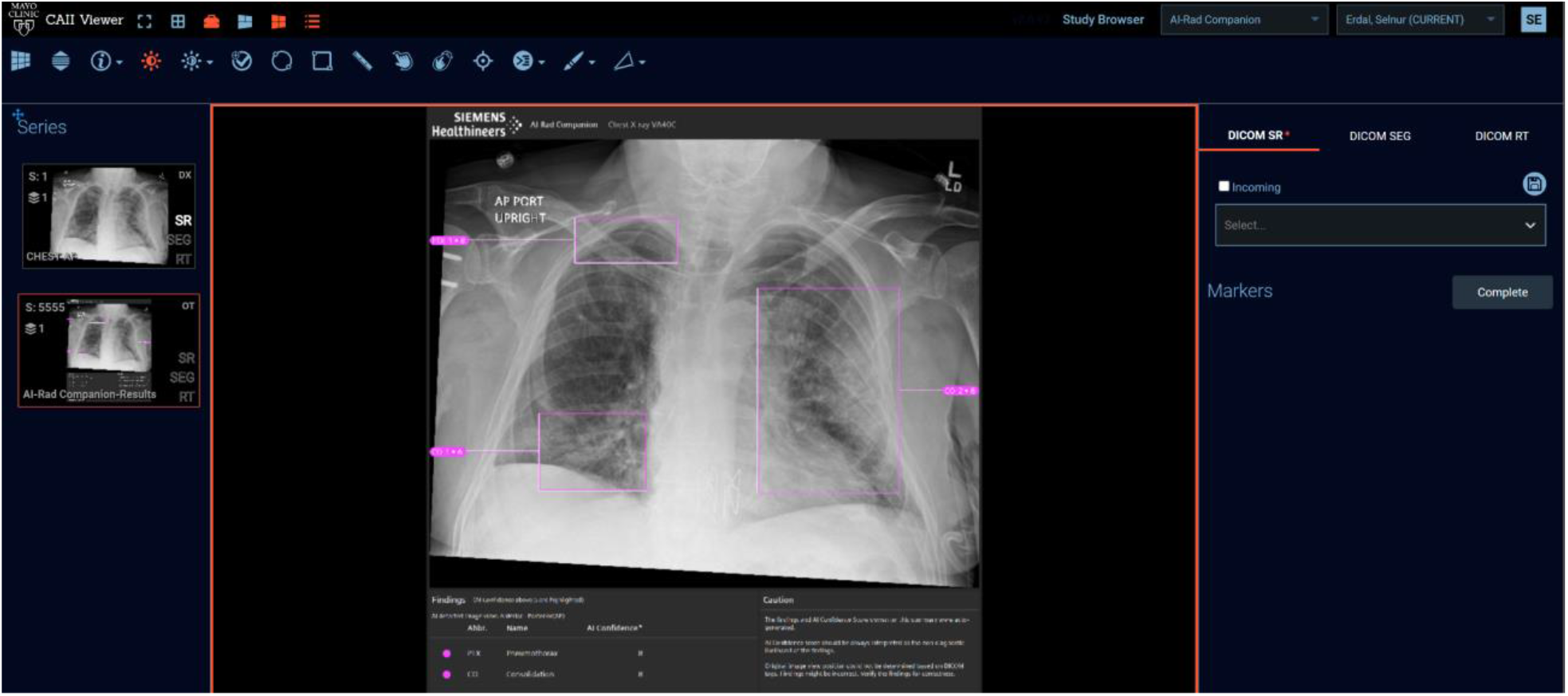
AIRC Chest X-Ray multi-finding case as displayed using the locally developed CAII Viewer. Like Figure 8a, the Secondary Captured annotations are not editable.

**Figure 9c:**
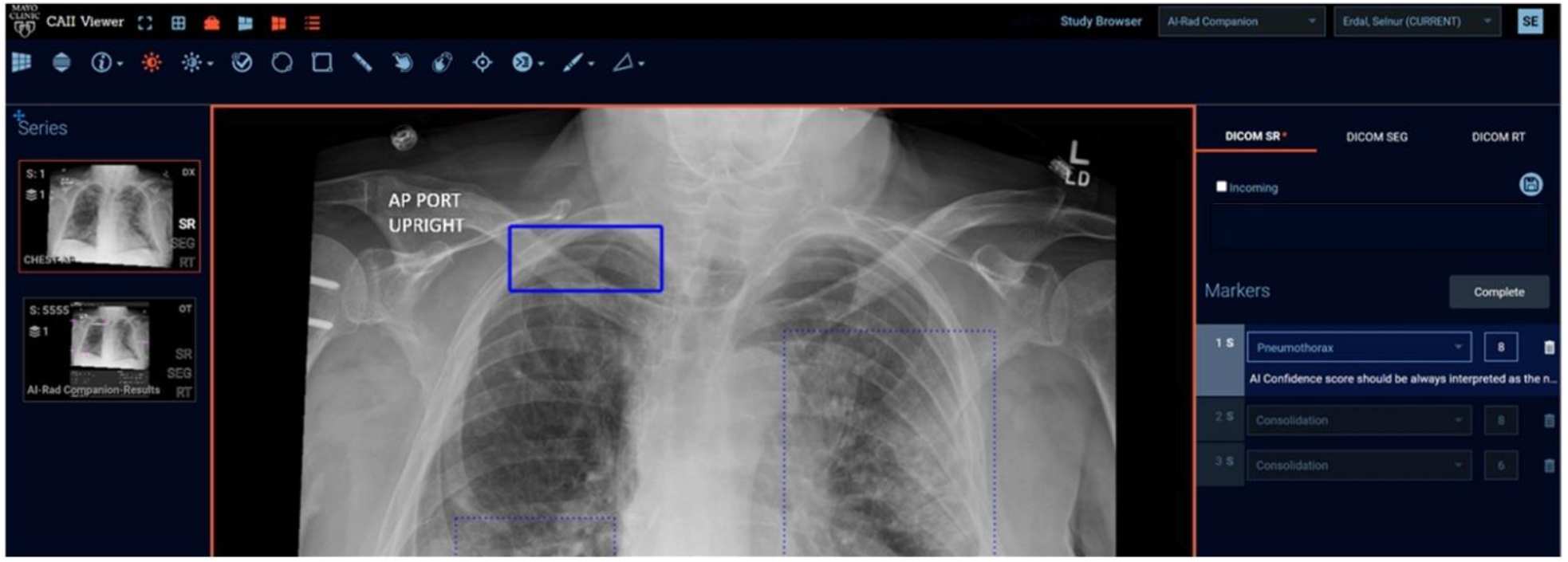
AIRC Chest X-Ray multi-finding case as displayed using the locally developed CAII Viewer (same patient as Figure 9b). The CAII Viewer displays DICOM SR TID1500-based annotations supporting post-inference adjudication workflow (accept/delete/edit options), as well as comprehensive, structured insights about these results (e.g., confidence score, comments extracted from the DICOM SR). A DICOM-SR-only workflow (e.g., not producing any DICOM Secondary Capture outputs) is possible in our environment.

**Figure 10:**
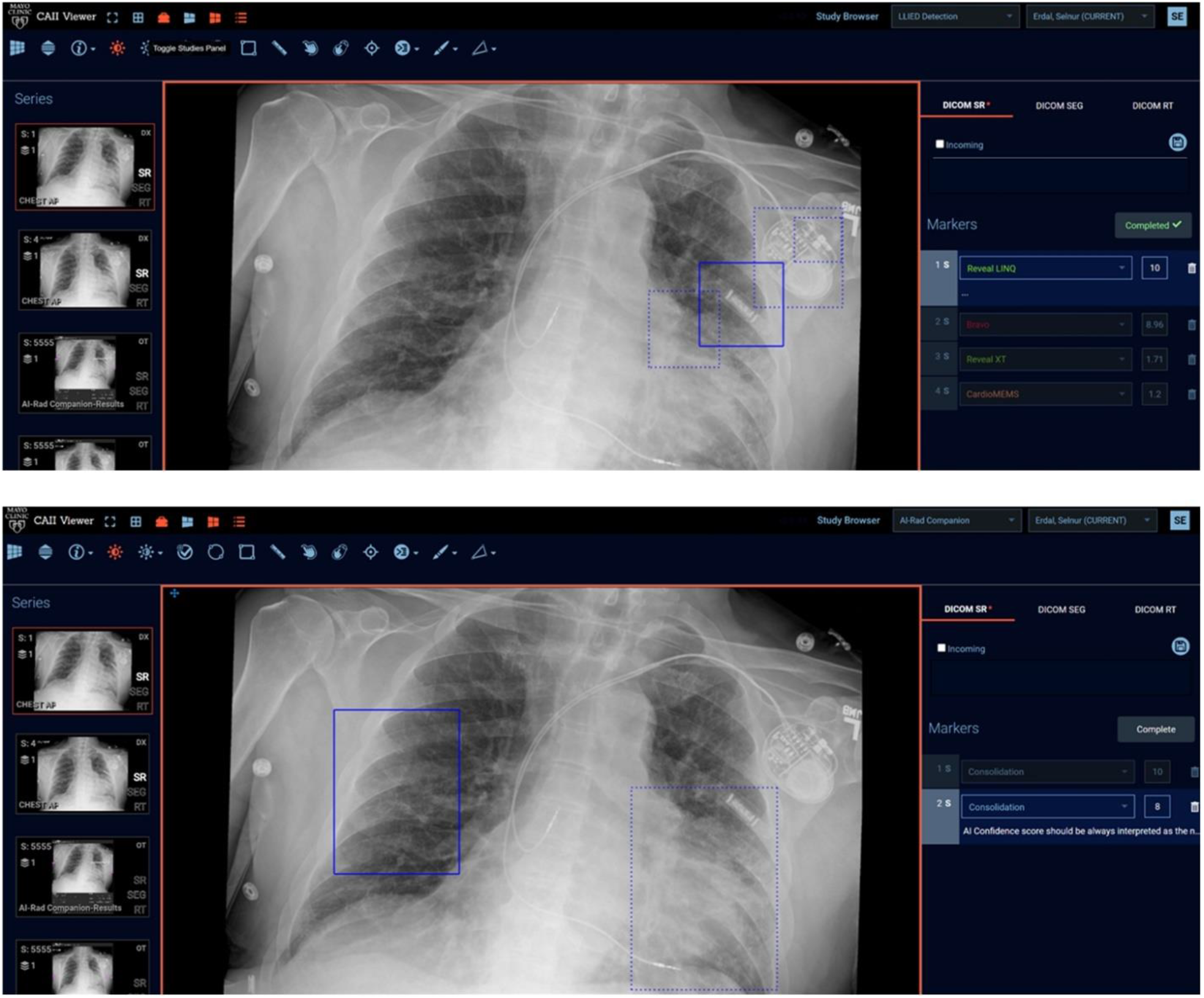
The standards-based CAII Viewer (exploiting formats like DICOM SR TID1500 according to IHE AIR) can allow the simultaneous display of inference results from different algorithmic processing of the same dataset. This is illustrated with the demonstrated results from AIRC Chest X-Ray (bilateral consolidation) and a locally developed algorithm for detection and identification of potentially MRI-unsafe leadless implanted electronic devices on Chest X-Rays.

## Conclusion

In this project, we have shown that algorithms, either vendor-based (e.g., Chest X-Ray AI solutions) or locally developed, can be simultaneously deployed and supported in parallel through a common standards-based architecture and workflow, even with complex operational requirements. Deployments of both types of solutions can co-exist in the same environment and provide concurrent AI support to end-users for a variety of tasks, including: image interpretation, inference result adjudication (for data collection, product improvement, or other purposes), or worklist management.

We successfully demonstrated that standards-based inference result production (e.g., as IHE-compliant DICOM SR TID1500), as well as result consumption by downstream applications, was possible. These capabilities could enable medical institutions to:

1. Gain control over AI results with which they interact (e.g., by providing edit/delete functionalities in appropriate viewing environments, therefore enabling adjudication of AI results).
2. View and interact with AI results from different sources (e.g., vendor-provided or locally developed algorithms)
3. Influence their reading workflow, if products cleared for that purpose are used (e.g., to flag time-sensitive cases in their worklist to promote timely reading and reporting). This could happen either by using middleware (parsing information from standard formats to transmit data points to a HIS/RIS worklist into a specific message format), as shown in this project, or by relying on existing functionalities of the HIS/RIS to ingest IHE-AIR-compliant datapoints.

With the implemented workflow based on several AI applications (different providers), we demonstrated that a viewer, such as our locally developed CAII Viewer, can deal with a variety of results (provided that both the viewer and the results adhere to certain standards). We illustrate two fundamental aspects of the IHE AIR profile, including that:

1. Following formats and result primitives recommended by IHE AIR can facilitate image display combined with baseline data-handling and presentation capabilities, making it “AI-ready” for supporting of a wide range of incoming results without the need for custom implementation;
2. It is beneficial for AI result generators (like the commercial and locally developed applications presented in this report) to support these data formats defined in IHE AIR, so that the produced results are compatible with a variety of displays and site workflows.

Using AI-results integration standards allowed us to have a successful implementation and timely installation of commercial products (e.g., AIRC) along with locally developed algorithms. Being able to track algorithm performances during testing, as well as being able to provide feedback on the results, helped us verify reliability and robustness prior to deployment. A formal evaluation is to be performed in a clinical trial setting and was not within the scope of this project. However, our initial impressions, which were very positive in terms of algorithm durability, were verifiable through performance data captured during this test phase.

## Data Availability

All data produced in the present work are contained in the manuscript.

